# Excess mortality in people living with HIV who received tuberculosis (TB) treatment with negative TB tests: a multinational study of data quality and validation

**DOI:** 10.64898/2026.06.24.26356079

**Authors:** Gustavo Amorim, Larissa Otero, Aihua Bian, Brenda Crabtree-Ramirez, Aggrey Semeere, Ran Tao, Stephany N. Duda, Beverly Musick, Constantin Yiannoutsos, Thomas Lumley, Pamela A. Shaw, Jessica L. Castillo, Lameck Diero, Timothy R. Sterling, Bryan E. Shepherd

## Abstract

**Background:** Observational studies typically use routinely collected (RC) data to investigate TB treatment outcomes, but these data may be error-prone. Using RC data together with validated data on a subsample, we explored the association of TB testing results with mortality and TB treatment outcomes in persons living with HIV (PLWH).

**Method:** We used RC data from two large HIV observational cohorts to evaluate the association of TB test result (positive/negative/unknown) at treatment start with death and the composite unfavorable outcome (treatment failure, loss-to-follow-up [LTFU], recurrence, or death) within 1.5 years of TB treatment start. We designed and implemented an optimal multi-wave validation study on a subsample of PLWH. We fitted logistic regression models using RC data only and combining them with the chart-validated data using a generalized raking approach, controlling for age, sex, body mass index (BMI), antiretroviral therapy, CD4 count, and region.

**Results:** RC data were extracted from 22,587 PLWH; 1122 were selected for validation, with 842 validated. We observed large discrepancies between validated and unvalidated data. We found that PLWH who started TB treatment with negative test results had higher odds of death compared to those who started treatment with a positive test: adjusted odds ratio (aOR)=1.22 (95% confidence interval [CI]=[1.03-1.45]) and aOR=3.94 (95% CI=[1.73-9.00]), using RC data only and RC plus validated data, respectively. Results for the composite outcome were inconclusive.

**Conclusion:** In PLWH treated for TB, bacteriological confirmation was associated with lower mortality. Data validation for observational research using RC clinical data care is needed.

## Introduction

Tuberculosis (TB) remains a major global health threat, accounting for over 1.24 million deaths in 2024^1^. Bacteriological confirmation plays a crucial role ensuring accurate identification of the disease, appropriate treatment, and testing for drug susceptibility.

This is particularly important for persons living with HIV (PLWH) who face higher risks of adverse TB outcomes and in whom other serious lung diseases are frequent, requiring early identification^2–5^.

Bacteriological confirmation, however, is not consistently done, reflecting implementation gaps and clinical or real-world constraints. At a global level, only 64% of pulmonary TB cases were bacteriologically confirmed in 2024^1^. The association between TB diagnosis and TB treatment outcomes, including mortality, has been explored in several studies^6–10^. Although evidence suggests PLWH who start TB treatment with negative TB test results have worse outcomes, these studies are limited by sample sizes and data quality concerns^11^.

Routinely collected (RC) surveillance and clinical data are crucial sources for programmatic and clinical decisions. While the World Health Organization (WHO) encourages integrated TB and HIV care, in many countries, they are treated separately with varying degrees of collaboration. TB and HIV diagnosis and treatment data often come from different sources: TB primary clinics where TB screening usually occurs and TB treatment is provided, TB specialized clinics where diagnosis of more complex cases (such as PLWH) is investigated, and from HIV primary or specialized clinics. In each of these settings, clinical, treatment, and laboratory data are frequently entered in separate registers^12,13^. Each of these data sources may have incomplete information, inconsistent measurements, or data entry errors that propagate through data integration, affecting disease monitoring, patient care, and research.

Given the central role of RC data in TB and HIV surveillance and observational research, validation of medical and laboratory records is essential to ensure data accuracy. Validating all records is impractical, as many countries have thousands of individuals co-infected with TB and HIV. In validation studies we address this constraint by selecting a subset of records for data validation and performing efficient statistical analyses that combine both validated and RC data^14,15^. Records are selected to maximize the precision of exposure-outcome associations. These approaches use multi-wave validation procedures that validate a certain number of records first, recalibrate the sampling procedure using the information learned in this first wave, and proceed to select another subset of records, an adaptive process that can be repeated over multiple waves^16^.

This study had two aims: 1) investigate the association between TB bacteriological test results and the risk of mortality and other poor TB treatment outcomes in a large and geographically diverse cohort of PLWH, and 2) determine RC data quality through extensive chart review. Using cutting-edge statistical techniques, we incorporate both the original RC data and validated data in our analyses, providing robust findings that correctly account for measurement errors.

## Methods

### Study cohort and eligibility criteria

We included data from 13 HIV clinical sites from 9 countries (Brazil, Chile, Haiti, Honduras, Mexico, Peru, Kenya, Tanzania, and Uganda) within two regions of the International epidemiology Databases to Evaluate AIDS (IeDEA) network: the Caribbean, Central and South America network for HIV Epidemiology (CCASAnet) and the East Africa region of IeDEA (EA-IeDEA). Participating sites are included in Supplementary Material.

RC electronic data from medical and laboratory records were assembled at each site and sent to CCASAnet and EA-IeDEA data coordinating centers at Vanderbilt University Medical Center and Indiana University, respectively, for processing, data quality checks, and harmonization. Institutional ethics review boards from participating sites and data coordinating centers approved the project (Vanderbilt IRB #060284 and #171353, and Indiana IRB #060478).

We extracted RC medical data from PLWH who had at least one visit to an HIV clinic between 2010-2018 and started TB treatment during the same period, and at least 450 days before database closure. PLWH were excluded if they had a prior TB diagnosis or treatment before TB treatment initiation.

### Study definitions

Electronic health records for eligible PLWH were tracked for 450 days (∼1.5 years) after their first TB treatment started. Outcomes were defined within this window. Outcome definitions are presented in **Table 1**, based on WHO definitions^13^. We created a composite outcome, *sustained TB treatment success*, to identify PLWH who were alive, still in HIV care, completed TB treatment within nine months of treatment start, and had no new TB diagnosis nor TB treatment initiation within 450 days of TB treatment start.

**Table 1.** Outcome definitions used in our study compared to definitions used by the World Health Organization (WHO).

| Outcome | Study definition | Comparison with WHO definition<br>(Restricted to TB treatment duration) |
| --- | --- | --- |
| Treatment failed | More than 270 days on TB treatment | Different definition. Defined as treatment that needed to be terminated or permanently changed. |
| Death | Death within 270 days after TB treatment start | Same definition |
| LTFU to TB treatment | Abandoning treatment or having treatment gaps greater than two consecutive months, during the first 270 days after TB treatment start | Same definition |
| LTFU to HIV care | Last HIV clinic visit within 450 days after TB treatment start and more than 1 year before database closing date | -- |
| Recurrence | New TB diagnosis after TB treatment completion, but before 450 days since TB treatment start | Same definition |
| Sustained TB treatment success | Treatment completed, alive, not LTFU to HIV, and no evidence of TB recurrence | Similar. Defined as successful treatment: either cured (with evidence of bacteriological response and no evidence of failure) or completed treatment (as recommended by national policy) |

Note: WHO outcome definition was extracted from the Tuberculosis Surveillance report, 2024 (Chapter 3)<sup>1</sup>
<sup>1</sup> 'Consolidated Guidance on Tuberculosis Data Generation and Use: Module 1: Tuberculosis Surveillance', n.d. <<https://www.who.int/publications/i/item/9789240075290>> [accessed 19 May 2026].

Our main exposure was TB test result at time of treatment initiation. PLWH could have received different types of TB tests: smear/microscopy, culture, Xpert MTB/RIF, and others not specified. For primary analysis we considered test results within 90 days before and 15 days after TB treatment initiation; test results outside this window were treated as *unknown*. In sensitivity analysis, we considered narrower (−30, +15 days) and wider windows (−90, +90 days) around treatment start. We defined PLWH as *positive* for TB if at least one test within this window was positive, irrespective of which test. PLWH were *negative* if all tests within this window were negative. PLWH without any test result within the window (either no test performed or with unavailable results) were categorized as *unknown*.

### Data Validation

To address the quality of RC data, we performed chart reviews (data validation) on a probabilistic sample of patient records. The Vanderbilt Data Coordinating Center randomly selected records for validation using a three-wave stratified sampling strategy that aimed to maximize precision of estimates of the adjusted log odds ratio between TB test result and the composite outcome of sustained TB treatment success, subject to a fixed chart-validation budget. Eighteen strata were defined by dividing the cohort into mutually exclusive groups based on three RC variables: sustained TB treatment outcome (successful/not successful), TB test result (positive/negative/unknown), and region (East Africa/Latin America/Haiti). Haiti was a unique stratum due to its large sample size relative to other Latin American countries. In wave 1, we used RC data to select an initial optimally allocated sample (n = 500 records), via Neyman allocation with influence functions^16–18^. In wave 2, we used validated data from wave 1 to select an additional 300 charts for review, and in wave 3, we used validated data from waves 1 and 2 to select 200 records for validation. Records in waves 2 and 3 were selected using an adaptive optimal design procedure^19^. Some charts could not be accessed; replacement records were selected for validation. More details are provided in Supplementary Material.

Data validation was performed at all 13 study sites by auditors with knowledge of clinical and data processes (e.g., research nurses and clinical fellows). Auditors attended a pre-audit training session. Sites received a list of record IDs. Auditors reviewed primary data sources and checked for consistency with the analysis dataset, focusing on specific variables including TB test result, elements used to define the composite outcome of sustained TB treatment success, and relevant covariates. Validation data were entered into REDCap^20,21^.

### Statistical Analysis

The primary analysis was pre-specified as multivariable logistic regression to investigate the association between TB test result and sustained TB treatment outcome, controlling for potential confounding variables measured at TB treatment initiation: age, sex, antiretroviral therapy (ART) use (naïve vs experienced), body mass index (BMI) and BMI’s interaction with age, CD4 count, year of TB treatment start, and region (East Africa, Latin America, Haiti). Additional analyses investigated associations between TB test result and individual elements of the composite outcome: death, loss to follow-up (LTFU; to TB treatment or HIV care), and treatment failure or recurrence among those who did not experience death or LTFU.

The primary analysis focused on exposure-outcome associations using generalized raking (GR) estimators^22^. GR estimators are calibrated inverse-probability weighted (IPW) estimators that fit regression models using validated data, weighting each subject by the inverse of their probability of being validated^23^. GR estimators calibrate the weights using RC data, allowing the RC information to contribute to the analysis, improving precision without additional assumptions over IPW estimators^24–26^.

Some records could not be validated (e.g., due to missing medical charts) while others could only be partially validated (e.g., only demographic variables). We defined a record as validated if the primary outcome was validated. We used logistic regression models to model the probability of being validated, given that the record was selected, adjusting for RC variables: stratification variables (composite TB treatment outcome, TB test result, and region), death and LTFU (either TB or HIV) status, as well as sex, age, and their interaction with the composite outcome. We assumed missing validation data as missing at random conditional on these covariates^27^. The weights in GR were computed as the inverse of the probability of being validated (known by design) and the probability of being validated given that the chart was selected for validation (estimated). More details are available in Supplementary Material.

Secondary analyses evaluated the association between TB test result and death using Kaplan-Meier curves and Cox proportional hazards models with unvalidated and validated data. Validated analysis used GR. All regression models accounted for missing covariates using multiple imputation with chained equations^28^ (50 imputations). Analyses were performed using R software^29^ and the *survey* package^30,31^.

## Results

### Overall characteristics and data validation

A total of 22,587 PLWH who initiated TB treatment were included: 19,085 (84%) from East Africa, 1370 (6%) from Latin America, and 2132 (9%) from Haiti. A total of 1122 participants were selected for chart review and 842 (75%) were fully validated.

Characteristics of records selected for validation are presented in **Supplementary Table S1**. In general, records that were unable to be validated came from PLWH who were more likely to be male, ART naïve, have negative TB tests, and be from Latin America.

**Table 2** shows characteristics of the study population, both based on unvalidated RC data and validated data weighted to be representative of the study population. The composition of the validated data, which was intentionally selected to overrepresent some groups of participants, is also shown. Overall, the median age in RC data was approximately 36 years, 50% were male, and 23% were ART naïve at TB treatment start.

**Table 2.** Description of the study population and discrepancies between routinely collected and chart-validated data.

| Characteristics | Unvalidated<br>RC data<br>(N = 22, 587) | Weighted<br>Estimates*<br>(N = 22, 587) | Chart-validated<br>(N = 842) | Percent<br>Error | Validation Metrics<br>(Cohen's Kappa $\kappa$, PPV/NPV or<br>Median Difference (Range)) |
| --- | --- | --- | --- | --- | --- |
| **TB test result** | | | | | $\kappa=0.62$ (95%CI: [0.59; 0.67]) |
| **Positive** | 10.5% | 19.3% | 37.5% | 16.7 | PPV=0.89, NPV=0.81 |
| **Negative** | 10.4% | 9.1% | 28.0% | 14.4 | PPV=0.70, NPV=0.93 |
| **Unknown** | 79.1% | 71.6% | 34.4% | 19.5 | PPV=0.69, NPV=0.88 |
| **Unsuccessful sustained TB**<br>**treatment outcome<sup>a</sup>** | 51.5% | 50.9% | 52.9% | 10.5 | PPV=0.94, NPV=0.85 |
| **Individual outcomes** |  |  |  |  |  |
| **Death<sup>b</sup>** | 16.3% | 20.9% | 18.1% | 6.1 | PPV=0.98, NPV=0.93 |
| **Treatment failure** | 11.0% | 5.5% | 6.4% | 25.1 | PPV=0.04, NPV=0.93 |
| **LTFU: TB** | 26.7% | 10.2% | 11.8% | 22.7 | PPV=0.20, NPV=0.90 |
| **LTFU: HIV** | 34.8% | 34.8% | 24.8% | 2.9 | PPV=0.95, NPV=0.98 |
| **Recurrence** | 3.0% | 1.9% | 2.1% | 2.0 | PPV=1.00, NPV=0.98 |
| **Male sex** | 50.3% | 50.5% | 61.2% | 0.2 | PPV=1.00, NPV=1.00 |
| **ART: Naive** | 23.0% | 26.3% | 37.0% | 5.9 | PPV=0.97, NPV=0.93 |
| **Age at treatment start (years)** | 35.7 (28.5; 43.7) | 35.1 [29.2; 44.7] | 34.9 (28.6, 43.1) | 7.8 | 1 (range -14, 6) |
| **CD4 count (cells/mm<sup>3</sup>)** | 165 (59; 346) | 149 [47; 318] | 136 (48, 307) | 12.2 | 57 [range -274, 644] |
| **BMI (kg/m<sup>2</sup>)** | 18.6 (16.4; 21.1) | 18.6 [17.1; 21.2] | 20.0 (17.6, 22.4) | 24.3 | 1.5 (range -32.5, 14.6) |
| **Weight (kg)** | 51.0 (44.0; 58.0) | 51.0 [42.0; 57.0] | 54.0 (45.8, 61.3) | 25.1 | 3.3 (range -30.8, 42) |
| **Height (m)** | 1.6 (1.6; 1.7) | 1.6 [1.6; 1.7] | 1.7 (1.6, 1.7) | 12.0 | 0.0 (range -0.2, 0.2) |

\*Weighted estimates are based the validated data ( $n = 842$ ), using calibrated inverse probability weights to make them representative of the full cohort.
<sup>a</sup>Composite unsuccessful sustained TB treatment outcome (treatment failure, incomplete TB treatment, loss to HIV care, death, or recurrence). <sup>b</sup>Death recorded within 1.5 years since treatment initiation.
Abbreviations: TB, tuberculosis; BMI, body mass index; LTFU, loss-to-follow-up; PPV, positive predictive value; NPV, negative predictive value; CI, confidence interval.

**Table 2** also shows error rates discovered in chart reviews and discrepancies between RC and validated data. In general, data discrepancies were observed across all variables, including exposures, outcomes, and demographic/laboratory variables. About 24% of validated records had different BMI values compared to RC, while CD4 differed in 12% of validated records, sometimes substantially (range of −274 to 644 cells/mm^3^). Death registration was more reliable, with 6% having errors and high positive (PPV) and negative (NPV) predicted values. RC TB tests results showed reasonable reliability (Cohen’s Kappa = 0.62): among test results classified as negative in RC data, 70% were confirmed as negative after chart validation (PPV = 0.70). Among positive tests in RC data, 89% were confirmed as positive (PPV = 0.89). Discrepancies between RC and chart validation varied substantially by region (**Supplementary Figure S1**).

Based on RC data, about 11% started TB treatment with a positive test, 10% started with a negative test, and 79% started with an unknown TB test result, with large variation by study region (**Table S2**). After validation, we estimated that approximately 19%, 9%, and 72% of patients started TB treatment with positive, negative, and unknown TB test results, respectively. **Table S3** in the Supplement shows the frequency of types of TB tests recorded in RC data. While most tests in East Africa were smear/microscopy, in Latin America and Haiti we also observed culture and Xpert MTB/RIF testing.

The frequency of unsuccessful TB treatment outcome was similar in RC data (52%) and weighted estimates after validation (51%). However, this composite outcome was misclassified for approximately 11% of validated participants, with varying individual outcome error rates. After accounting for data validation, we estimated that 21% died, 11% and 35% were LTFU to TB treatment or to HIV care, respectively, 6% had treatment failure, and 2% had TB recurrence (**Table 2**).

### Associations between test results and treatment outcomes

Associations between baseline TB test results and TB treatment outcomes are shown in **Table 3**. There was little evidence of an association between baseline TB test and the composite outcome of unsuccessful sustained TB treatment, although those with unknown TB test results had an increased odds of unsuccessful TB treatment outcome in the adjusted analysis with the unvalidated RC data. Estimates were generally similar, although with wider confidence intervals, in weighted analyses that incorporated chart-validated records.

**Table 3.** Odds ratios (and 95% confidence intervals) measuring associations between TB treatment test result and study outcomes in unadjusted and adjusted analyses, using unvalidated routinely collected (RC) data and using weighted analyses combining RC and chart-validated data via generalized raking. Adjusted analyses included age, sex, antiretroviral therapy use, body mass index, CD4 count, year of TB treatment start, and region.

|  | Unvalidated RC data |  | Weighted analysis |  |
| --- | --- | --- | --- | --- |
|  | OR (95% CI) | p-value | OR (95% CI) | p-value |
| **Unsuccessful sustained TB treatment outcome** |  |  |  |  |
| *Unadjusted* |  |  |  |  |
| Positive | Reference |  | Reference |  |
| Negative | 0.93 (0.83; 1.05) | 0.260 | 1.03 (0.54; 1.96) | 0.930 |
| Unknown | 0.94 (0.86; 1.02) | 0.134 | 0.83 (0.50; 1.39) | 0.482 |
| *Adjusted* |  |  |  |  |
| Positive | Reference |  | Reference |  |
| Negative | 0.97 (0.85; 1.10) | 0.594 | 0.75 (0.39; 1.45) | 0.400 |
| Unknown | 1.19 (1.07; 1.33) | <0.001 | 1.02 (0.56; 1.87) | 0.948 |
| **Death** |  |  |  |  |
| *Unadjusted* |  |  |  |  |
| Positive | Reference |  | Reference |  |
| Negative | 1.14 (0.97; 1.34) | 0.113 | 2.29 (0.99; 5.32) | 0.054 |
| Unknown | 1.12 (0.99; 1.27) | 0.074 | 1.66 (0.84; 3.27) | 0.146 |
| *Adjusted* |  |  |  |  |
| Positive | Reference |  | Reference |  |
| Negative | 1.22 (1.03; 1.45) | 0.025 | 3.94 (1.73; 9.00) | 0.001 |
| Unknown | 1.11 (0.95; 1.29) | 0.204 | 1.70 (0.78; 3.72) | 0.183 |
| **LTFU (either TB or HIV care)** |  |  |  |  |
| *Unadjusted* |  |  |  |  |
| Positive | Reference |  | Reference |  |
| Negative | 1.07 (0.95; 1.21) | 0.292 | 1.27 (0.65; 2.50) | 0.488 |
| Unknown | 1.19 (1.08; 1.30) | <0.001 | 1.31 (0.75; 2.27) | 0.343 |
| *Adjusted* |  |  |  |  |
| Positive | Reference |  | Reference |  |
| Negative | 0.88 (0.77; 1.00) | 0.046 | 1.01 (0.53; 1.92) | 0.981 |
| Unknown | 1.20 (1.08; 1.34) | <0.001 | 1.28 (0.68; 2.24) | 0.453 |
| **Failure/Recurrence** |  |  |  |  |
| *Unadjusted* |  |  |  |  |
| Positive | Reference |  | Reference |  |
| Negative | 0.68 (0.54; 0.85) | 0.001 | 1.44 (0.52; 4.04) | 0.481 |
| Unknown | 0.45 (0.38; 0.53) | <0.001 | 0.26 (0.09; 0.76) | 0.014 |
| *Adjusted* |  |  |  |  |
| Positive | Reference |  | Reference |  |
| Negative | 0.85 (0.67; 1.08) | 0.181 | 0.61 (0.24; 1.59) | 0.315 |
| Unknown | 0.74 (0.61; 0.90) | 0.003 | 0.41 (0.13; 1.34) | 0.140 |

Abbreviations: TB, tuberculosis; BMI, body mass index; LTFU, loss-to-follow-up; OR, odds ratio; CI, confidence interval.

In contrast, both unadjusted and adjusted models showed strong associations between negative TB test results and increased odds of dying. Compared to PLWH with a positive TB test, those with a negative TB test at baseline were at increased odds of dying: unadjusted (OR) and adjusted odds ratios (aOR) of 1.14 (95% confidence interval [CI] = [0.97-1.34]) and 1.22 (95% CI = [1.03-1.45], respectively, in analyses with unvalidated RC data. We observed stronger associations with death after incorporating the validated data: OR = 2.29 (95% CI = [0.99-5.32] and aOR = 3.94 (95% CI = [1.73-9.00]).

Similar findings were observed in time-to-death analyses (**Supplementary Table S4**). Using RC data, we observed 23% increased risk of death for those with a negative TB vs. positive TB test (adjusted hazard ratio (aHR) = 1.23, 95% CI = [1.05-1.44]). This hazard was higher after incorporating chart-validated data: aHR = 3.13 (95% CI = [1.68-5.84]). Kaplan-Meier curves are displayed in **Supplementary Figure S2**.

Although RC data analysis suggested that PLWH who started TB treatment with unknown TB test results were at higher odds of LTFU, this association did not hold after incorporating validation data. Similarly, no associations were seen for the combined failure and recurrence analyses, although confidence interval for GR analyses were wide.

Results were largely similar in sensitivity analyses that varied the window for eligible TB treatment test results (**Supplementary Tables S5 and S6**) and that extended time on treatment to 365 days to define TB treatment failure (**Supplementary Tables S7 and S8**).

## Discussion

We designed a large, multi-wave validation study in a multinational TB/HIV cohort to evaluate the impact of starting TB treatment without bacteriological confirmation on treatment outcomes and to understand the role of RC data quality in these analyses. We used modern robust statistical methods to design an optimal validation study and to efficiently combine validated and unvalidated data for inference.

Although we did not observe a statistically significant association between TB test results and the composite unfavorable TB treatment outcome, we identified a strong and consistent association between TB tests results and mortality. PLWH who started TB treatment with negative TB test result had higher odds of death, compared to PLWH with bacteriological confirmed TB, in both unadjusted and adjusted analyses. This effect was even greater using generalized raking estimators that incorporated both unvalidated and validated data.

We estimated that fewer than 20% of PLWH started TB treatment with positive test results. The large proportion without a test result may reflect different scenarios: unavailability of testing, inability to obtain a sample, insufficient test sensitivity to detect TB, delays in receiving test confirmation, or an unidentified alternative clinical etiology. There was also large variability in the number of TB tests performed across regions.

Errors in RC data were frequent and present in all three regions. Death had the lowest error rate, while LTFU, an outcome more prone to interpretation and site heterogeneity, had one of the highest. The high frequency of errors may be partly explained by the fragmentation of TB/HIV care. PLWH access different health services throughout the continuum of care — peripheral and primary care clinics, hospital outpatient and inpatient departments, and private and public facilities — resulting in multiple data sources from HIV clinics, TB clinics, laboratories, and death registers. Data may be captured directly in electronic systems (or still first on paper in some sites), but despite recent expansion of digital registration in TB and HIV programs, these systems rarely communicate. For example, TB and HIV clinics often rely on staff manually searching electronic laboratory databases for test results and re-entering them into their own systems. TB and HIV programs often do not harmonize data on numbers of coinfected cases and treatment outcomes. Some differences are also due to follow-up demands of each disease: TB patients require at least nine months follow up while HIV demands lifelong follow-up.

This study had several limitations. We were unable to exactly match WHO definitions for TB outcomes; our proxy composite outcome was imperfect, challenging interpretation.

Some records selected for chart validation could not be found. Although modeled the probability of being fully validated given a record was selected for validation, those which were not validated may have systematically differed from those validated. We could not distinguish between new TB cases or recurrent infections, limiting discussion about TB recurrence or treatment failure. We did not distinguish between persons who started empiric anti-TB therapy pending microbiology results who subsequently had microbiologic confirmation versus those who did not.

Nevertheless, our study had many strengths. We used large, heterogeneous datasets, from nine countries in three regions with high burdens of HIV and/or TB. We used a novel, optimal statistical design targeting who were more informative for the research question for data validation. We used robust generalized raking estimators that incorporated both validated and unvalidated data, making full use of information available without imposing strict modeling assumptions. Sensitivity analyses supported our findings, highlighting robustness of results.

By carefully designing and analyzing data from a large validation study, our findings are aligned with previous studies, that the availability of confirmation tests in the context of TB diagnosis could potentially improve outcomes and reduce mortality among PLWH and TB^6,32^. Despite access to TB diagnostic testing, alternative or concomitant etiologies may remain unrecognized. Consequently, patients may receive incomplete or suboptimal management that does not fully address the underlying disease processes, potentially contributing to adverse clinical outcomes and a poorer prognosis. Second, the data quality of routine electronic health records for TB and HIV needs to be urgently addressed for valid inference in programmatic decision making and research studies, improving data flows and interoperating electronic systems, and implementing efficient data validation practices.

## Data Availability

All data produced in the present study are available upon reasonable request to the authors

## Footnotes

1 ‘Consolidated Guidance on Tuberculosis Data Generation and Use: Module 1: Tuberculosis Surveillance’, n.d. <https://www.who.int/publications/i/item/9789240075290> [accessed 19 May 2026].

## Notes

### Competing Interest Statement

The authors have declared no competing interest.

### Author Declarations

Ethics committee/IRB Vanderbilt University Medical Center and Indiana University gave ethical approval for this work

### Funding

- National Institutes of Health: U01AI069923, U01AI069911, R37AI131771
- National Heart, Lung, and Blood Institute Division of Intramural Research: R01HL149422

