## Supplementary Material for "Excess mortality in people living with HIV who received tuberculosis (TB) treatment with negative TB tests: a multinational study of data quality and validation"

**Supplementary Table S1.** Comparison of routinely collected data among all PLWH selected for chart validation by whether chart validation was completed.

|  | *Unable to validate or*  *only partially validated*  *(N = 280)* | *Fully validated*  *(N = 842)* | p-value |
| --- | --- | --- | --- |
| TB test result: |  |  | <0.001 |
| Positive | 41 (14.6%) | 223 (26.5%) |  |
| Negative | 117 (41.8%) | 283 (33.6%) |  |
| Unknown | 122 (43.6%) | 336 (39.9%) |  |
| Unsuccessful sustained TB treatment outcome | 139 (49.6%) | 405 (48.1%) | 0.705 |
| Individual outcomes |  |  |  |
| Death | 56 (20.0%) | 105 (12.5%) | 0.003 |
| Treatment failure | 55 (19.6%) | 171 (20.3%) | 0.877 |
| LTFU: TB | 35 (12.5%) | 152 (18.1%) | 0.039 |
| LTFU: HIV | 73 (26.1%) | 207 (24.6%) | 0.676 |
| Recurrence | 0 (0.00%) | 1 (0.12%) | >0.999 |
| Male sex | 192 (68.6%) | 518 (61.5%) | 0.040 |
| Age at treatment start | 34.6 [27.6;42.2] | 34.8 [28.5;43.1] | 0.391 |
| CD4 count (cells/mm^3^) | 110 [36.0;295] | 137 [49.0;310] | 0.146 |
| ART status at TB diagnosis: Naive | 73 (30.2%) | 237 (33.8%) | 0.343 |
| BMI (kg/m^2^) | 20.4 [17.7;23.8] | 19.6 [17.2;22.3] | 0.022 |
| Weight (kg) | 55.8 [48.0;65.0] | 53.0 [45.0;61.5] | 0.03 |
| Height (m) | 1.66 [1.59;1.70] | 1.66 [1.57;1.71] | 0.473 |
| Region: |  |  | <0.001 |
| EA-IeDEA | 84 (30.0%) | 383 (45.5%) |  |
| CCASAnet | 158 (56.4%) | 383 (45.5%) |  |
| Haiti | 38 (13.6%) | 76 (9.03%) |  |

Abbreviations: PLWH, persons living with HIV; TB, tuberculosis; BMI, body mass index; LTFU, loss-to-follow-up.

**Supplementary Table S2.** Overall description of the study population and weighted estimates obtained by combining the routinely collected and chart-validated data, stratified by study region.

| *Characteristics* | *Unvalidated routinely RC data*  *(*$\boldsymbol{N=22,587}$*)* | | | | *Chart-validated (weighted)*  *(*$\boldsymbol{N=842}$*)* | | |
| --- | --- | --- | --- | --- | --- | --- | --- |
|  | **EA-IeDEA**  *(*$\boldsymbol{N=19,085}$*)* | **CCASAnet**  *(*$\boldsymbol{N=1370}$*)* | **Haiti**  *(*$\boldsymbol{N=2132}$*)* | **EA-IeDEA**  *(*$\boldsymbol{N=383}$*)* | | **CCASAnet**  *(*$\boldsymbol{N=383}$*)* | **Haiti**  *(*$\boldsymbol{N=76}$*)* |
| Unsuccessful sustained TB treatment outcome | 9544 (50.0%) | 914 (66.7%) | 1169 (54.8%) | 7564 (48.5%) | | 899 (65.6%) | 1150 (54.0%) |
| TB test result |  |  |  |  | |  |  |
| Positive | 1138 (6.0%) | 588 (42.9%) | 493 (23.1%) | 2408 (15.5%) | | 824 (60.1%) | 628 (29.5%) |
| Negative | 982 (5.2%) | 287 (20.9%) | 891 (41.8%) | 702 (4.5%) | | 297 (21.6%) | 924 (43.3%) |
| Unknown | 16965 (88.9%) | 495 (36.1%) | 748 (35.1%) | 12474 (80.0%) | | 250 (18.2%) | 580 (27.2%) |
| Age at treatment start | 35.9 [28.7;43.6] | 34.0 [27.9;42.7] | 35.3 [27.4;44.8] | 36.0 [29.2, 42.1] | | 34.7 [28.6, 43.2] | 35.0 [26.3, 45.7] |
| Male sex | 9172 (48.1%) | 1101 (80.4%) | 1082 (50.8%) | 7609 (48.8%) | | 1051 (76.8%) | 1022 (47.9) |
| Height (m) | 1.6 [1.6;1.7] | 1.7 [1.6;1.7] | 1.6 [1.5;1.7] | 1.6 [1.6, 1.7] | | 1.7 [1.6, 1.7] | 1.6 [1.5, 1.7] |
| Weight (kg) at TB treatment initiation | 51.0 [44.0;58.0] | 60.0 [53.0;67.0] | 51.0 [44.0;58.0] | 50.0 [42.0, 57.0] | | 57.0 [50.2, 63.0] | 50.0 [43.0, 56.3] |
| BMI (kg/m^2^) | 18.5 [16.4;20.9] | 22.1 [19.6;24.5] | 18.0 [15.9;20.3] | 18.5 [16.8, 20.8] | | 20.7 [18.5, 22.6] | 17.6 [16.2, 20.5] |
| CD4 count | 163 [58;344] | 108 [44;242] | 264 [117;466] | 134 [39, 306] | | 116 [45, 256] | 320 [175, 458] |
| ART status at TB diagnosis: Naive | 1872 (22.3%) | 602 (43.9%) | 261 (12.2%) | 2209 (26.5%) | | 689 (52.5%) | 221 (10.4%) |
| TB treatment outcomes^1^ |  |  |  |  | |  |  |
| Death^b^ | 3197 (16.8%) | 266 (19.4%) | 224 (10.5%) | 3265 (21.0%) | | 338 (24.7%) | 475 (22.3%) |
| Treatment failure | 1798 (9.4%) | 503 (36.7%) | 179 (8.4%) | 511 (3.3%) | | 121 (8.8%) | 510 (23.9%) |
| LTFU: TB | 5063 (26.5%) | 200 (14.6%) | 0 (0.0%) | 1163 (7.5%) | | 261 (19.1%) | 613 (28.8%) |
| LTFU: HIV | 6475 (33.9%) | 340 (24.8%) | 1048 (49.2%) | 5562 (35.7%) | | 313 (22.8%) | 750 (35.2%) |
| Recurrence | 44 (0.2%) | 57 (4.2%) | 36 (1.7%) | 198 (1.3%) | | 35 (2.6%) | 181 (8.5%) |

Abbreviations: TB, tuberculosis; BMI, body mass index; LTFU, loss-to-follow-up.

**Supplementary Table S3**. Total number of TB tests performed available in the routinely collected data, stratified by region. Patients could have repeated testing with the same or different types of tests.

| *Type of TB test and test results* | *EA-IeDEA*  *(N = 2185)* | *CCASAnet*  *(N = 1980)* | *Haiti*  *(N = 2563)* |
| --- | --- | --- | --- |
| Smear/microscopy |  |  |  |
| Positive | 1011 (46.3%) | 425 (21.5%) | 373 (14.6%) |
| Negative | 956 (43.8%) | 271 (13.7%) | 993 (38.7%) |
| Total | **1967 (90.0%)** | **696 (35.1%)** | **1366 (53.3%)** |
| Culture |  |  |  |
| Positive | 9 (0.4%) | 325 (16.4%) | 402 (15.7%) |
| Negative | 3 (0.1%) | 212 (10.7%) | 132 (5.2%) |
| Total | **12 (0.5%)** | **537 (27.1%)** | **534 (20.8%)** |
| GeneXpert |  |  |  |
| Positive | 149 (6.8%) | 62 (3.1%) | 325 (12.7%) |
| Negative | 57 (2.6%) | 49 (2.5%) | 338 (13.2%) |
| Total | **206 (9.5%)** | **112 (5.7%)** | **663 (25.9%)** |
| PCR |  |  |  |
| Positive | 0 (0.0%) | 5 (0.3%) | 0 (0.0%) |
| Negative | 0 (0.0%) | 28 (1.4%) | 0 (0.0%) |
| Total | **0** | **33 (1.7%)** | **0** |
| Other TB test |  |  |  |
| Positive | 0 (0.0%) | 236 (11.9%) | 0 (0.0%) |
| Negative | 0 (0.0%) | 366 (18.5%) | 0 (0.0%) |
| Total | **0** | **602 (30.4%)** | **0** |

Abbreviations: TB, tuberculosis

**Supplementary Table S4**. Hazard ratios (and 95% confidence intervals) measuring associations between TB treatment test result and time-to-death in unadjusted and adjusted region-stratified Cox regression models, using unvalidated routinely collected (RC) data and using weighted analyses combining RC and chart-validated data via generalized raking. Adjusted analyses included age, sex, antiretroviral therapy use, body mass index, CD4 count, year of TB treatment start, and region.

|  | Unvalidated RC data | | Weighted analysis | |
| --- | --- | --- | --- | --- |
|  | **HR (95% CI)** | **p-value** | **HR (95% CI)** | **p-value** |
| *Unadjusted* |  |  |  |  |
| Positive | Reference | | Reference | |
| Negative | 1.29 (1.10; 1.50) | <0.001 | 2.36 (1.18; 4.71) | 0.016 |
| Unknown | 1.06 (0.94; 1.20) | 0.346 | 1.65 (0.91; 3.00) | 0.103 |
| *Adjusted* |  | |  | |
| Positive | Reference | | Reference | |
| Negative | 1.23 (1.05; 1.44) | 0.013 | 3.13 (1.68; 5.84) | <0.001 |
| Unknown | 1.14 (0.99; 1.33) | 0.079 | 1.79 (0.96; 3.53) | 0.070 |

Abbreviations: HR, hazard ratio; CI, confidence interval.

**Supplementary Table S5**. Sensitivity analysis using a narrower window of (-30, +15) days around TB treatment start date to compute test results. Results are displayed as odds ratios (and 95% confidence intervals), measuring associations between TB treatment test result defined within this narrower window and study outcomes in unadjusted and adjusted analyses, using unvalidated routinely collected (RC) data and using weighted analyses combining RC and chart-validated data via generalized raking. Adjusted analyses included age, sex, antiretroviral therapy use, body mass index, CD4 count, year of TB treatment start, and region.

|  | Unvalidated RC data | | Weighted analysis | |
| --- | --- | --- | --- | --- |
|  | **OR (95% CI)** | **p-value** | **OR (95% CI)** | **p-value** |
| Unsuccessful sustained TB treatment outcome |  |  |  |  |
| *Unadjusted* |  |  |  |  |
| Positive | Reference | | Reference | |
| Negative | 0.95 (0.84; 1.07) | 0.354 | 1.11 (0.59; 2.10) | 0.755 |
| Unknown | 1.06 (0.97; 1.16) | 0.208 | 0.93 (0.56; 1.54) | 0.771 |
| *Adjusted* |  |  |  |  |
| Positive | Reference | | Reference | |
| Negative | 0.87 (0.76; 0.98) | 0.023 | 0.89 (0.46; 1.72) | 0.721 |
| Unknown | 1.22 (1.10; 1.36) | <0.001 | 1.04 (0.56; 1.93) | 0.907 |
| Death |  |  |  |  |
| *Unadjusted* |  |  |  |  |
| Positive | Reference | | Reference | |
| Negative | 1.14 (0.97; 1.34) | 0.113 | 2.65 (1.14; 6.14) | 0.023 |
| Unknown | 1.12 (0.99; 1.27) | 0.074 | 1.72 (0.88; 3.36) | 0.111 |
| *Adjusted* |  |  |  |  |
| Positive | Reference | | Reference | |
| Negative | 1.22 (1.03; 1.45) | 0.024 | 4.37 (1.79; 10.69) | 0.001 |
| Unknown | 1.11 (0.95; 1.30) | 0.186 | 1.63 (0.74; 3.59) | 0.228 |
| LTFU (either TB or HIV care) |  |  |  |  |
| *Unadjusted* |  |  |  |  |
| Positive | Reference | | Reference | |
| Negative | 1.02 (0.91; 1.15) | 0.710 | 1.22 (0.63; 2.39) | 0.554 |
| Unknown | 1.15 (1.05; 1.25) | 0.003 | 1.40 (0.81; 2.42) | 0.231 |
| *Adjusted* |  |  |  |  |
| Positive | Reference | | Reference | |
| Negative | 0.88 (0.77; 1.00) | 0.046 | 1.01 (0.53; 1.92) | 0.981 |
| Unknown | 1.20 (1.08; 1.34) | <0.001 | 1.28 (0.68; 2.40) | 0.453 |
| Failure/Recurrence |  |  |  |  |
| *Unadjusted* |  |  |  |  |
| Positive | Reference | | Reference | |
| Negative | 0.68 (0.54; 0.85) | 0.001 | 1.44 (0.52; 4.00) | 0.481 |
| Unknown | 0.45 (0.38; 0.53) | <0.001 | 0.26 (0.09; 0.76) | 0.014 |
| *Adjusted* |  |  |  |  |
| Positive | Reference | | Reference | |
| Negative | 0.85 (0.67; 1.08) | 0.181 | 0.61 (0.24; 1.59) | 0.315 |
| Unknown | 0.74 (0.61; 0.90) | 0.003 | 0.41 (0.13; 1.34) | 0.140 |

Abbreviations: HR, hazard ratio; CI, confidence interval.

**Supplementary Table S6**. Sensitivity analysis using a narrower window of (-90, +90) days around TB treatment start date to compute test results. Results are displayed as odds ratios (and 95% confidence intervals), measuring associations between TB treatment test result defined within this narrower window and study outcomes in unadjusted and adjusted analyses, using unvalidated routinely collected (RC) data and using weighted analyses combining RC and chart-validated data via generalized raking. Adjusted analyses included age, sex, antiretroviral therapy use, body mass index, CD4 count, year of TB treatment start, and region.

|  | Unvalidated RC data | | Weighted analysis | |
| --- | --- | --- | --- | --- |
|  | **OR (95% CI)** | **p-value** | **OR (95% CI)** | **p-value** |
| Unsuccessful sustained TB treatment outcome |  |  |  |  |
| *Unadjusted* |  |  |  |  |
| Positive | Reference | | Reference | |
| Negative | 0.93 (0.83; 1.04) | 0.177 | 0.84 (0.46; 1.52) | 0.559 |
| Unknown | 0.92 (0.84; 1.00) | 0.052 | 0.87 (0.53; 1.43) | 0.585 |
| *Adjusted* |  |  |  |  |
| Positive | Reference | | Reference | |
| Negative | 0.96 (0.86; 1.09) | 0.545 | 0.53 (0.29; 0.98) | 0.042 |
| Unknown | 1.16 (1.04; 1.28) | 0.006 | 1.06 (0.58; 1.94) | 0.861 |
| Death |  |  |  |  |
| *Unadjusted* |  |  |  |  |
| Positive | Reference | | Reference | |
| Negative | 1.20 (1.03; 1.40) | 0.021 | 2.01 (0.89; 4.56) | 0.094 |
| Unknown | 1.11 (0.99; 1.25) | 0.079 | 1.68 (0.87; 3.22) | 0.121 |
| *Adjusted* |  |  |  |  |
| Positive | Reference | | Reference | |
| Negative | 1.26 (1.07; 1.48) | 0.005 | 2.48 (1.13; 5.44) | 0.024 |
| Unknown | 1.10 (0.96; 1.27) | 0.18 | 1.66 (0.80; 3.44) | 0.176 |
| LTFU (either TB or HIV care) |  |  |  |  |
| *Unadjusted* |  |  |  |  |
| Positive | Reference | | Reference | |
| Negative | 1.06 (0.94; 1.18) | 0.362 | 1.07 (0.56; 2.05) | 0.840 |
| Unknown | 1.17 (1.07; 1.28) | <0.001 | 1.35 (0.79; 2.30) | 0.268 |
| *Adjusted* |  |  |  |  |
| Positive | Reference | | Reference | |
| Negative | 0.96 (0.85; 1.08) | 0.51 | 0.63 (0.33; 1.17) | 0.145 |
| Unknown | 1.25 (1.13; 1.39) | <0.001 | 1.27 (0.68; 2.37) | 0.463 |
| Failure/Recurrence |  |  |  |  |
| *Unadjusted* |  |  |  |  |
| Positive | Reference | | Reference | |
| Negative | 0.76 (0.66; 0.89) | <0.001 | 1.13 (0.43; 3.02) | 0.801 |
| Unknown | 0.44 (0.39; 0.49) | <0.001 | 0.26 (0.09; 0.74) | 0.012 |
| *Adjusted* |  |  |  |  |
| Positive | Reference | | Reference | |
| Negative | 1.00 (0.85; 1.18) | 0.994 | 0.47 (0.15; 1.42) | 0.18 |
| Unknown | 0.68 (0.59; 0.79) | <0.001 | 0.47 (0.14; 1.62) | 0.232 |

Abbreviations: HR, hazard ratio; CI, confidence interval.

**Supplementary Table S7.** Descriptive analysis of unsuccessful sustained TB outcome and treatment failure, after defining treatment failure as prolonged treatments (over 365 days) or with two-month gaps within TB treatment.

| *Outcomes* | *Unvalidated RC data*  *(*$\boldsymbol{N=22,587}$*)* | | | | *Chart-validated (weighted)*  *(*$\boldsymbol{N=842}$*)* | | |
| --- | --- | --- | --- | --- | --- | --- | --- |
|  | **EA-IeDEA**  **(N = 19085)** | **CCASAnet**  **(N =1370)** | **Haiti**  **(N = 2132)** | **EA-IeDEA**  **(N = 383)** | | **CCASAnet**  **(N = 239)** | **Haiti**  **(N = 88)** |
| Unsuccessful sustained TB treatment outcome | 9011 (47.2%) | 669 (48.8%) | 1205 (56.5%) | 7564 (48.5%) | | 899 (65.6%) | 1150 (54.0%) |
| TB treatment failure | 771 (4.04%) | 206 (15.0%) | 103 (4.83%) | 511 (3.3%) | | 121 (8.8%) | 510 (23.9%) |

Abbreviations: TB, tuberculosis.

**Supplementary Table S8.** Odds ratios (and 95% confidence intervals) to assess the effect of treatment effect on unsuccessful sustained TB treatment effect, after defining treatment failure as prolonged treatments (over 365 days) or with two-month gaps within TB treatment, obtained from unvalidated routinely collected (RC) data and using weighted analyses combining RC and chart-validated data via generalized raking. Adjusted analyses included age, sex, antiretroviral therapy use, body mass index, CD4 count, year of TB treatment start, and region.

|  | Unvalidated RC data | | Weighted analysis | |
| --- | --- | --- | --- | --- |
|  | **OR (95% CI)** | **p-value** | **OR (95% CI)** | **p-value** |
| *Unadjusted* |  |  |  |  |
| Positive | Reference | | Reference | |
| Negative | 0.95 (0.84; 1.07) | 0.354 | 1.11 (0.59; 2.10) | 0.755 |
| Unknown | 1.06 (0.97; 1.16) | 0.208 | 0.93 (0.56; 1.54) | 0.771 |
| *Adjusted* |  | |  | |
| Positive | Reference | | Reference | |
| Negative | 0.87 (0.76; 0.98) | 0.023 | 0.89 (0.46; 1.72) | 0.721 |
| Unknown | 1.22 (1.10; 1.36) | <0.001 | 1.04 (0.60; 1.93) | 0.907 |

Abbreviations: OR, odds ratio; CI, confidence interval.

**Supplementary Figure S1.** Percent error for all main variables by region: EA-IeDEA (EA); CCASAnet (CN), and Haiti.

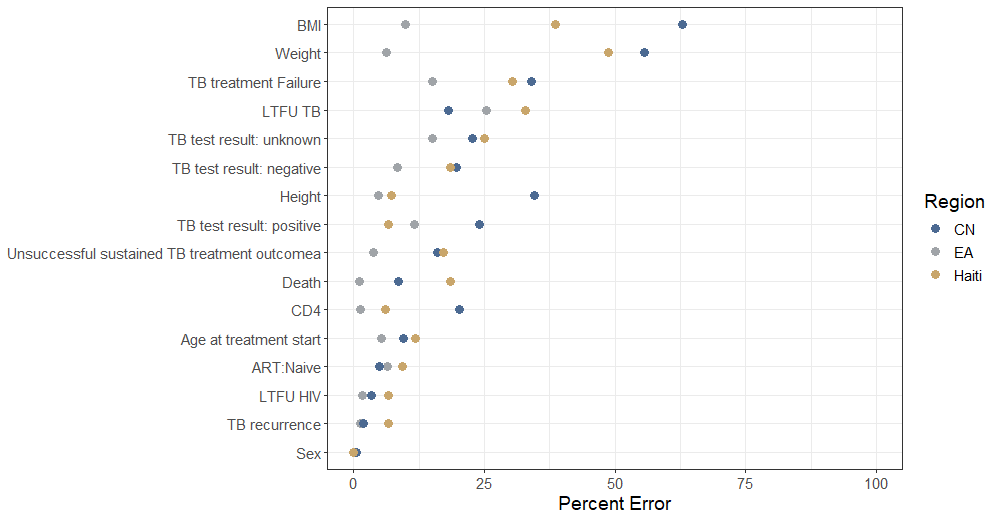

**Supplementary Figure S2.** Kaplan-Meier curves showing associations between TB test results and death. Left panel are results using unvalidated data; right panel show estimates using generalized raking with validated data. 95% confidence intervals are included.

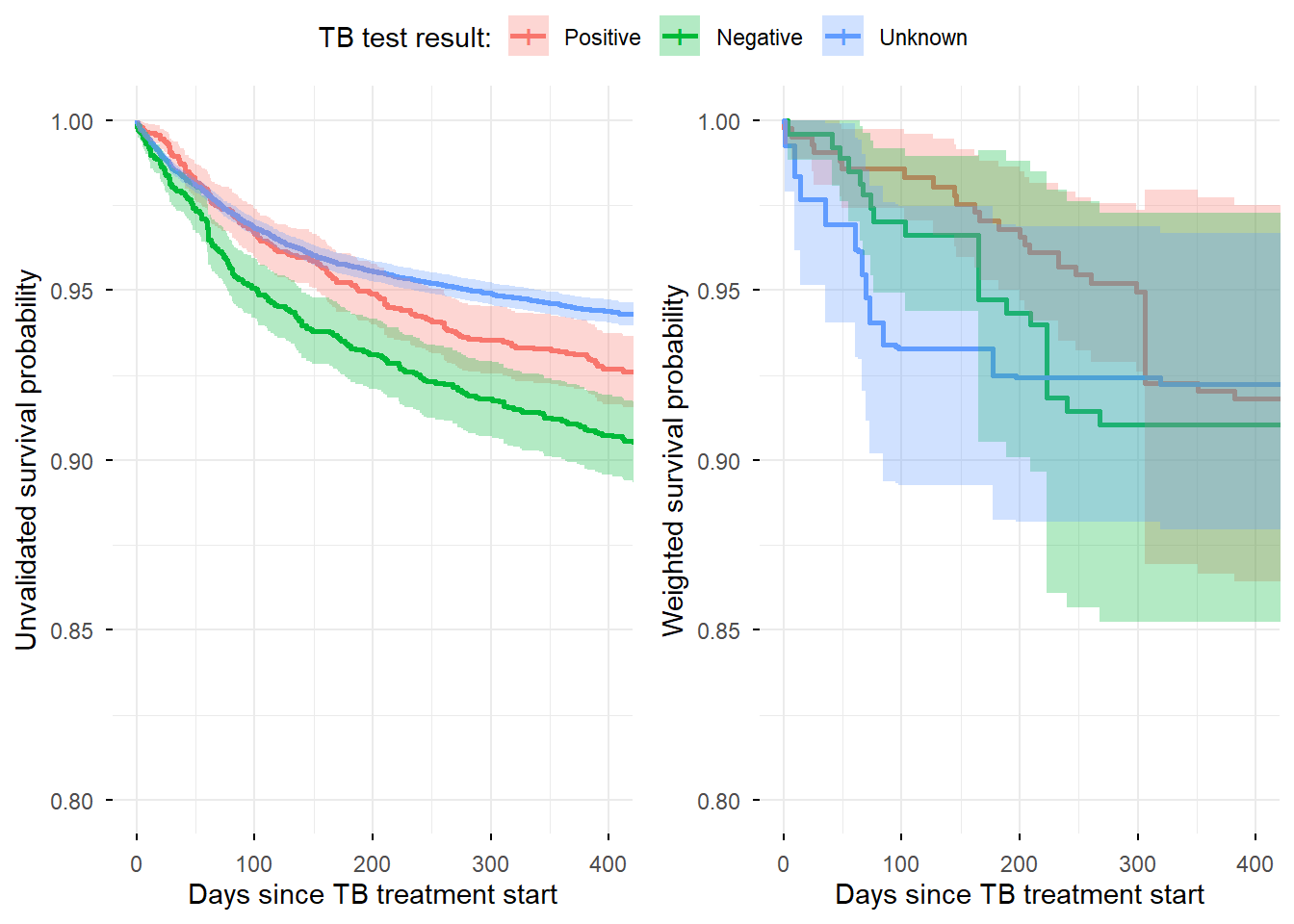

**Participating Sites**

Participating sites from the Caribbean, Central and South America network for HIV Epidemiology (CCASAnet) were

- Instituto Nacional de Infectologia - Evandro Chagas, Fundação Oswaldo Cruz, Rio de Janeiro, Brazil
- Fundacion Arriaran, Santiago-Chile
- Groupe Haitien d’Etude du Sarcome de Kaposi et des Infections Opportunistes (GHESKIO), Port-au-Prince, Haiti
- Instituto Hondureño de Seguridad Social and Hospital Escuela, Tegucigalpa, Honduras
- Instituto Nacional de Ciencias Médicas y Nutrición Salvador Zubirán, Mexico City, Mexico
- Instituto de Medicina Tropical Alexander von Humboldt, Lima, Perú

Participating sites from the East African region of the International epidemiology Databases to Evaluate AIDS (EA-IeDEA) were

- Academic Model Providing Access to Healthcare (AMPATH), Eldoret, Kenya
- Family AIDS Care & Education Services (FACES), Kisumu, Kenya
- Infectious Disease Institute (IDI), Kampala, Uganda
- Masaka Regional Hospital HIV Care Clinic, Masaka City, Uganda
- Morogoro Regional Hospital CTC, Morogoro Region, Tanzania
- Rakai Health Sciences Program, Rakai District, Uganda
- Tumbi Regional Referral Hospital CTC, Kibaha Town, Pwani Region, Tanzania

**Rationale for selecting records for chart validation**

We initially aimed to validate a total of $n$ records, split into multiple waves. In wave 1, the only information to guide sampling is the error-prone routinely collected (RC) data. The simplest approach is selecting records at random for chart validation. Although this leads to a representative subset of records, it is inefficient as it ignores the rich phase-1 data. Instead, we stratified the cohort into 18 mutually exclusive strata based on three RC variables: TB treatment outcome (sustained/no sustained), TB test result (positive/negative/unknown), and region (EA‑IeDEA, CCASAnet, Haiti). We decided on a stratified sampling scheme, with random sampling within strata.

We next used Neyman allocation to determine the initial number of records within each stratum to be validated. Neyman allocation increases statistical precision by sampling more heavily from strata many records and with high variability. This variability is measured as the standard deviation of the influence function; a function that measures the impact of removing a single record on an estimated parameter. Formally, Neyman allocation is given by

$$n_{k}=n\frac{N_{k}s_{k}}{\sum_{k} N_{k}s_{k}},$$

where $N_{k}$ is the total number of records in stratum $k$ and $n_{k}$ is the number of records from the *k*th to be sampled for validation and $s_{k}$ is the within stratum standard deviation for stratum *k*.

For wave 2, we used the optimal allocation procedure described in Lotspeich et. al. (2024). This approach uses the validated data from wave 1 to estimate misclassification rates and identify strata with higher error rates to be oversampled so that the asymptotic variance of the maximum likelihood estimator (MLE) for the exposure-outcome association is reduced. It works by searching for the optimal allocation of $n_{1},\ldots, n_{K}$ using a grid search under a budget and stratum size constraints. The variance of the exposure-outcome estimator is computed for each $\left( n_{1},\ldots, n_{K} \right)$ configuration and the allocation with smallest variance is selected. To reduce computational burden, an adaptive grid search was originally implemented, starting with a coarse grid to identify the direction of the gradient descent, narrowing down to a fine grid as the range of possible values decreases.

The optimal allocation algorithm oversamples from strata with large proportions of errors, maximizing the efficiency gained by the MLE. When applying the optimal design in wave 2, we noticed that its allocation would target very heavily from CCASAnet and Haiti strata. We were concerned that this could be reflecting small numbers of validated records rather than the true underlying error rates. To avoid overreliance on a few validated strata, we decided to select fewer records in wave 2 for validation and leave the remaining pre-specified number of records to be selected in a subsequent wave 3.

Once the wave‑2 validations were completed, we updated our estimates and re-run the optimal allocation algorithm to select the remaining number of records for validation. Because of logistical problems in one site in CCASAnet (delayed IRB approval), we were forced to remove data from this specific site and re-run the grid search algorithm. The resulting wave‑3 sample completed the total validation target and provided the final set of validated records used for SMLE and the alternative estimators.

**Generalized raking estimator**

Generalized raking (GR) is an estimator that builds upon inverse probability weighting (IPW) to construct more efficient estimators, i.e., statistical estimators with smaller variance. It was first introduced in the survey sampling literature, but has been increasingly applied in biostatistics.

In standard IPW, each participant selected into phase-2 (e.g., for chart validation) receives a weight that is the inverse of its probability of being sample. That is, if we define this sampling probability as $\pi_{i}$, where *i* ($i=1, \ldots, n$) denotes the *i*th patient selected for chart validation, the *i*th participant receives the weight ${w_{i}=1/ \pi}_{i}$. IPW estimators are well-known to be consistent, but inefficient, i.e., they estimate the quantity that would have been estimated if had chart-reviewed all patients, but the estimator will be imprecise, resulting in wide confidence intervals.

Generalized raking estimators make use of auxiliary information available in the full cohort to adjust the sampling weights $w=\left( w_{1},\ldots, w_{n} \right)$. It replaces the IPW weights with a set of calibrated weights $\tilde{w}_{i}$ such that the *calibration constraint* $\sum_{i=1}^{N} R_{i}\tilde{w}_{i}A_{i}=\sum_{i=1}^{N} A_{i}$ are always satisfied. This constraint adjusts the IPW weights in such a way that the weighted quantity $\tilde{w}_{i}A_{i}$, observed in the validation sample matches the total observed in the study cohort (routinely collected data, our phase-1 data).

The optimal auxiliary variable (i.e., the one that reduces the variance of the parameter estimate of interest) depends on the problem, whether we are estimating a population total or a regression parameter. Here we are interested in the latter. Breslow et. al.^[1]^ showed that the optimal variable is the *influence function*, a mathematically constructed object that quantifies how the estimated parameters changes if we remove one subject from the analysis at a time.

In our settings, the probability of being validated is a product of two probabilities: the probability of being selected for chart validation times the probability of being validated given that the record was selected for validation. The first probability is known by design and is a function of the stratification variables: composite TB treatment outcome, TB test result, and region. The second is unknown and needs to be estimated. We fitted a logistic regression model, using the following variables available in the routinely collected data as predictors: stratification variable (composite TB treatment outcome, TB test result, and region), death and LTFU (either TB or HIV) status, as well as sex, age, and their interaction with the composite outcome. The IPW weights used in generalized raking was the inverse of the product of these two probabilities.

The calibration variable was obtained from the routinely collected data. We fitted the outcome model using the error-prone phase-1 data and extract the influence functions associated with the parameters of interest (TB test results: negative and unknown; positive was the baseline). We repeated this procedure for every imputation (50 in total) and average the influence functions across the imputations. The averaged influence functions were used to calibrated the weights, following the steps described above.

Generalized raking can be directly used in R statistical software. It is implemented in the *survey* R package ^[2]^.

1. Norman E. Breslow and others, ‘Improved Horvitz-Thompson Estimation of Model Parameters from Two-Phase Stratified Samples: Applications in Epidemiology’, *Statistics in Biosciences*, 1.1 (2009), p. 32, doi:10.1007/s12561-009-9001-6.
2. Thomas Lumley, ‘Survey: Analysis of Complex Survey Samples’, *R Package Version 4.4*, n.d.
